# Automated Quantification of Decreased FAF in Stargardt Disease: Validation of a Novel Method Compared to Manual Grading Standards

**DOI:** 10.1101/2025.07.07.25330927

**Authors:** Mohamed I. Ahmed, Hikmet Yucel, Rubbia Afridi, Thales A.C. de Guimarães, Isabel Sendino-Tenorio, Nam V. Nguyen, Kiran Romaisa, Sidrah Khan, Ufaq Khan, Syeda Sharaf un Nisa, Mauro Campigotto, Amir Hariri, Michel Michaelides, Hendrik P.N. Scholl, Nathan Mata, Quan D. Nguyen, Yasir J. Sepah

## Abstract

**Purpose:** To evaluate the repeatability and reproducibility of a novel automated method compared with manual segmentation for measuring decreased autofluorescence (DAF) and definitely decreased autofluorescence (DDAF) in fundus autofluorescence (FAF) images of patients with Stargardt disease.

**Design:** Cross-sectional reproducibility and agreement study.

**Participants:** A total of 316 eyes from 158 genetically confirmed Stargardt patients were analyzed. For intra-grader repeatability, 114 FAF images were reassessed in a masked, repeated-measures design.

**Methods:** DAF and DDAF lesion areas were independently quantified by five certified graders using either manual delineation with Heidelberg RegionFinder or a threshold-based automated algorithm. Agreement and repeatability were assessed using intraclass correlation coefficients (ICC), standard error of measurement (SEM), minimal detectable change (MDC), Lin’s concordance correlation coefficient (CCC), Bland–Altman plots, and Passing–Bablok regression. Both raw and square-root-transformed lesion areas were evaluated.

**Main Outcome Measures:** Repeatability (intra-grader ICC, SEM, MDC), reproducibility (inter-grader ICC), and agreement (CCC, bias in regression analysis) between and within manual and automated methods.

**Results:** The automated method achieved excellent intra-grader repeatability for both DAF and DDAF (ICCs ≥0.988, SEM ≤0.71 mm², MDC ≤1.98 mm²), with minimal operator influence. Manual measurements showed variable repeatability (DAF ICCs 0.909–0.974; DDAF ICCs as low as 0.837), with square-root transformation reducing SEM and MDC. Inter-grader reproducibility was highest for automated methods (ICC = 0.989–0.992), whereas manual methods ranged from 0.764–0.939 (raw) and 0.867–0.922 (transformed). Cross-method agreement was strong (CCC = 0.91–0.96), though minor proportional and constant bias was observed in raw DAF data.

**Conclusions:** The automated approach provides near-perfect repeatability and high agreement with manual grading, offering a scalable, objective alternative for quantifying hypo-autofluorescent lesions in Stargardt disease. Manual methods are generally reliable but more variable, especially for DDAF, and benefit from square-root transformation.

## Introduction

Fundus autofluorescence (FAF) imaging is a critical tool for monitoring retinal degenerations, including Stargardt disease (STGD1), due to its capacity to visualize lipofuscin distribution in the retinal pigment epithelium (RPE) in vivo – frequently before more prominent changes appear in the fundus^1^. STGD1, caused by ABCA4 mutations, leads to lipofuscin accumulation and eventually to progressive RPE and photoreceptor atrophy. On FAF, affected areas typically appear as dark regions of decreased autofluorescence (DAF), which are often described as well-demarcated in the literature^2^. However, in clinical practice, lesion boundaries can vary considerably, and in many cases, the demarcation is indistinct, particularly at the lesion margins. Initial quantification of DAF relied on manual planimetry, as shown in a longitudinal study by Fujinami et al. (2013), which tracked atrophic area growth over nearly a decade^3^. Although effective, manual tracing is laborious and subject to reader variability. To standardize assessments, tools like Heidelberg’s RegionFinder introduced semiautomated segmentation using seed-and-grow algorithms and adjustable thresholds, but considerable human involvement remained necessary^4^.

The ProgStar study, a multicenter effort led by Scholl and colleagues, formalized DAF measurement protocols for clinical trials^5^. FAF images were graded for “definitely decreased” (DDAF; ≥90% black relative to the optic disc) and “questionably decreased” (QDAF; 50–90%) autofluorescence. RegionFinder was used to segment lesions, with lesion area quantified for both DDAF and QDAF alone and DDAF+QDAF combined. This protocol improved consistency and became the standard for evaluating STGD1 progression^6^.

Despite these advances, variability in lesion delineation remains a challenge. ProgStar addressed this through dual-reader workflows and adjudication of non-consensus cases. Still, subtle differences arise, especially at lesion borders or in differentiating DDAF from QDAF. Although Georgiou et al. (2020) reported excellent intraclass correlation coefficients (ICCs) of 0.995 (intra) and 0.987 (inter) for total DAF area, significant non-consensus was found in around 42% of cases, mostly due to disagreements on whether an area has 50-90% or 90-100% DAF leading the investigators to decide to only compare measurements of DAF^4^. Factors like image quality, illumination, and lesion edge gradation contribute to measurement variability.

STGD1 progression varies with disease stage. In early stages, atrophy progresses slowly (∼0.05– 0.1 mm²/year), but in advanced stages, expansion can exceed 4 mm²/year^3^. ProgStar’s 12- and 24-month findings confirmed average lesion growth of ∼0.5–0.8 mm²/year^5, 7^. Such small margins of change require higher precision in the measuring procedure. While these studies validated DAF as a reproducible metric, they also highlighted the burden of human grading.

To address this, researchers have turned to automation. Early thresholding methods^8^ lacked sensitivity for STGD1’s heterogeneity. More recently, Zhao et al. (2022) developed a deep learning model for FAF lesion segmentation, achieving high agreement with expert graders (Dice score ∼0.90 for DDAF). Performance for QDAF was lower (∼0.55), reflecting inherent ambiguity^9^.

In summary, although FAF-based DAF quantification is central to STGD1 research, it is constrained by manual workflows and inter-reader variability. While emerging automated techniques offer promising solutions, this study aims to build on these efforts, proposing a novel, reproducible approach for quantifying DAF to support future clinical applications.

## Methods

### Study Design and Participants

This observational study was conducted to assess the intra- and inter-grader repeatability and agreement of two lesion endpoints, Definitely Decreased Autofluorescence (DDAF) and overall Decreased Autofluorescence (DAF), using two distinct measurement approaches: a traditional semi-automated method based on a seed-and-growth algorithm (referred to as the manual method in this manuscript), and a novel automated method employing a threshold-based approach. The concordance between these two methods was evaluated to determine the degree of alignment between the automated system and established manual practices. A total of 316 eyes from 158 subjects diagnosed with Stargardt disease were retrospectively included. Data were curated from fundus autofluorescence (FAF) images acquired during the screening visits of the LBS-008-CT03 Clinical Trial (DRAGON) [ClinicalTrials.gov ID: NCT05244304]. The study adhered to the tenets of the Declaration of Helsinki and received approval from the local institutional review board. Informed consent was obtained from all participants prior to inclusion.

### Measurement Protocol

Five certified graders (RA, TG, IST, RK, NN) independently assessed the reduced autofluorescence measurements on the same set of images. Each grader performed a second round of measurements on 114 randomly selected images after a washout period of at least one week between sessions to minimize recall bias. Two distinct decreased autofluorescence endpoints were evaluated based on the definitions of the ProgStar study standard of DDAF and DAF *(combined QDAF+DDAF)*.

For further analysis, each endpoint was also transformed using the square root function, yielding the transformed variables sqDAF and sqDDAF. This transformation, which is commonly employed in the analysis of similar lesion types such as geographic atrophy^10, 11^, was applied to mitigate heteroscedasticity and reduce the influence of outliers, thereby improving the statistical robustness of subsequent reliability assessments.

### Measurement Methods

Three graders (IST, RK, NN) performed manual delineation of lesion boundaries using a traditional semi-automated approach, referred to as the manual method in this manuscript (Region Finder, Heidelberg Engineering). This method relies on a seed-and-growth algorithm in which lesion boundaries are automatically generated by the software following grader input. Graders initiate the process by selecting a seed point within the atrophic area, and the software automatically delineates lesion boundaries, allowing manual adjustments to parameters such as “growth power” to refine segmentation. However, since graders retain full control over the thresholding parameters, particularly the “growth power,” this directly influences the extent of lesion boundary propagation.

Two graders (RA, TG) employed a novel automated image analysis algorithm designed to enable high-throughput and reproducible quantification of autofluorescence lesion areas. The method requires the grader to manually identify the foveal center and the optic disc, as well as to provide anatomical reference points used to calibrate a uniform spatial scale. This calibration facilitates pixel-wise classification based on the characteristics of local neighborhoods relative to pre-specified, tunable intensity thresholds. The algorithm incorporates configurable parameters, including the ability to set a minimum lesion size threshold for inclusion in the aggregate lesion area, defined as ≥0.5 mm² in this study. It also allows restriction and stratifications of measurements to predefined regions of interest within the fundus. In this study, all measurements were limited to the central 6 mm of the fundus. The algorithm also supports vessel masking to exclude retinal vasculature from analysis, thereby minimizing the risk of overestimating lesion size. While the algorithm is capable of operating across multiple threshold levels, including those capturing the hyperautofluorescent spectrum, only two threshold levels, selected to reflect grading standards for Stargardt disease, were used in this study.

### Statistical Analysis

All statistical analyses were conducted using industry-standard software, including IBM SPSS Statistics (Version 30). Descriptive statistics (mean, standard deviation, and range) were computed for each lesion endpoint (DAF and DDAF), both in their original form and after square-root transformation, to account for potential skewness and heteroscedasticity.

Square-root transformation was applied to all DAF and DDAF values to stabilize variance and reduce skewness, as detailed in the Data Transformation and Rationale section. All reliability and agreement metrics are reported for both raw and transformed data.

Intra-grader repeatability was assessed using the Intraclass Correlation Coefficient (ICC) under a two-way mixed-effects model with absolute agreement definition (single rater, single measurement). For each grader, ICCs were calculated separately for raw and transformed data. To quantify absolute measurement error, the Standard Error of Measurement (SEM) was derived using the formula:

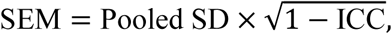

where pooled standard deviation was computed as:

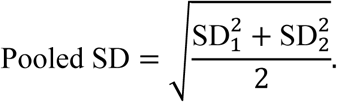

The Minimal Detectable Change (MDC), also referred to as the Coefficient of Repeatability (CR), was calculated as:

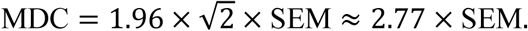

Inter-grader repeatability (within-method reliability) was assessed using the same ICC framework for each pairwise comparison among graders operating under the same method (automated or manual). For each pair, both single-measure and average-measure ICCs were computed, along with SEM and MDC. The effect of square-root transformation was evaluated by comparing reliability metrics before and after transformation.

Agreement between methods (automated vs. manual) was assessed using Lin’s Concordance Correlation Coefficient (CCC) to quantify overall concordance and Passing–Bablok regression to detect proportional and constant bias. CCC was computed manually using its standard definition, incorporating Pearson’s correlation, standard deviations, and mean differences. Passing–Bablok regression provided estimates of slope and intercept to assess systematic deviations from the identity line. For each endpoint and transformation state, the CCC, regression slope, and intercept were reported.

Bland–Altman analysis was used to visualize intra- and inter-grader agreement within each method. Separate Bland–Altman plots were constructed for each pairwise comparison (raw and transformed), stratified by method and lesion type. To accommodate the number of comparisons (n = 20 for intra-grader and n = 16 for inter-grader), composite figures were generated to display representative plots for automated and manual graders across both lesion phenotypes.

Throughout the analysis, special attention was given to evaluating the effect of square-root transformation on measurement precision and agreement. Comparisons were made across lesion types (DAF vs. DDAF), measurement methods (automated vs. manual), and data scales (raw vs. transformed) to understand the contribution of each factor to reliability and concordance.

### Data Transformation and Rationale

All DAF and DDAF measurements were initially inspected for normality and skewness. Histograms, Shapiro–Wilk tests, and calculations of skewness indicated that the data were substantially right-skewed, with a small number of markedly larger lesions inflating the mean and variance. Such heavy skew can undermine parametric assumptions in reliability and agreement analyses. Accordingly, a square-root transformation *(√x)* was applied to all raw measurements (DAF, DDAF) to stabilize variance and reduce outlier influence.

Nonetheless, both raw and transformed analyses are presented in this study to allow the clinical community to interpret the lesion measurements using the original scale. Specifically, we report inter- and intra-grader reliability and method-comparison results (e.g., ICC, Bland–Altman limits, Concordance Correlation Coefficients) on the untransformed (raw) data *and* on the square-root transformed data. This approach ensures that readers can (1) gauge performance under standard clinical units and (2) appreciate any improvements in statistical robustness afforded by the transformation.

## Results

FAF images from a total of 316 eyes of 158 subjects were assessed in the study, with a mean age of 15.4 ± 2.25 years (range, 12–21 years). The cohort included 91 males (57.6%) and 67 females (42.4%). Most of participants identified as Asian (58.9%), followed by Caucasian (34.2%), with a small proportion identifying as Other (5.1%) or declining to report race (1.9%) (**Table 1**).

**Table 1.**
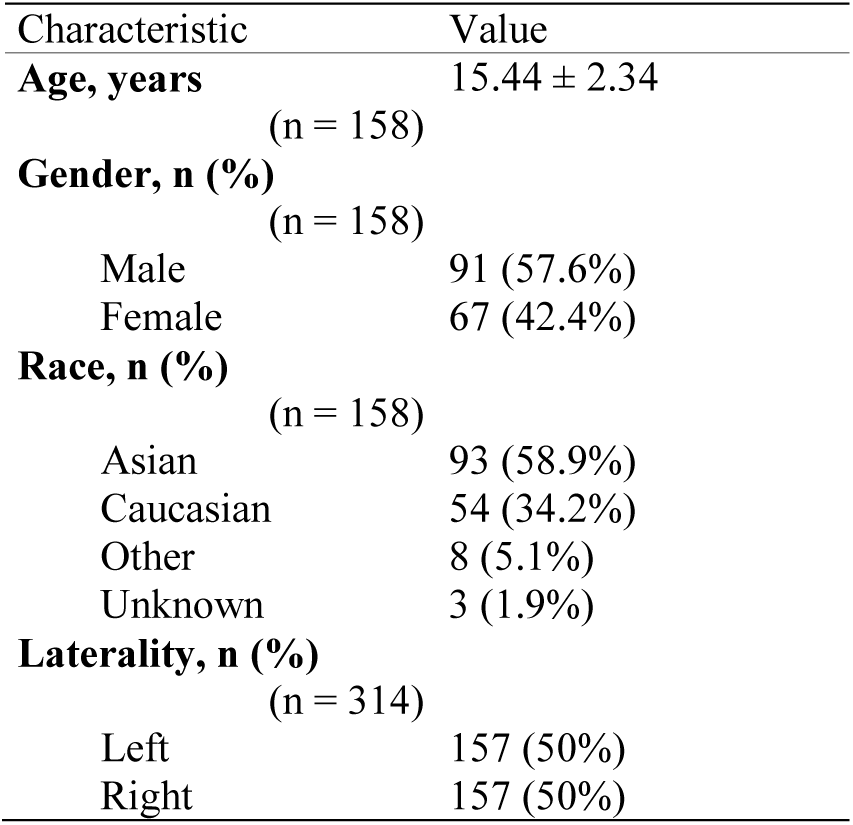
Baseline demographic and clinical characteristics of study participants. Values are reported as mean ± standard deviation for continuous variables and as counts with percentages for categorical variables. BCVA = best-corrected visual acuity.

| Characteristic | Value |
| --- | --- |
| **Age, years** | 15.44 ± 2.34 |
| (n = 158) |  |
| **Gender, n (%)** |  |
| (n = 158) |  |
| Male | 91 (57.6%) |
| Female | 67 (42.4%) |
| **Race, n (%)** |  |
| (n = 158) |  |
| Asian | 93 (58.9%) |
| Caucasian | 54 (34.2%) |
| Other | 8 (5.1%) |
| Unknown | 3 (1.9%) |
| **Laterality, n (%)** |  |
| (n = 314) |  |
| Left | 157 (50%) |
| Right | 157 (50%) |

Images from two eyes (2 subjects) were deemed ungradable by all graders due to insufficient quality to perform any type of analysis. Additional 12 images were deemed ungradable for manual grading by at least two of the three graders in the manual graders group. **Table 2** summarizes the computed values for each grader and endpoint, both on the raw scale (DAF and DDAF) and after square root transformation (sqDAF and sqDDAF). **Figure 1** shows representative examples of manual and automated delineation of DAF and DDAF lesions across a range of lesion sizes and configurations.

**Figure 1.**
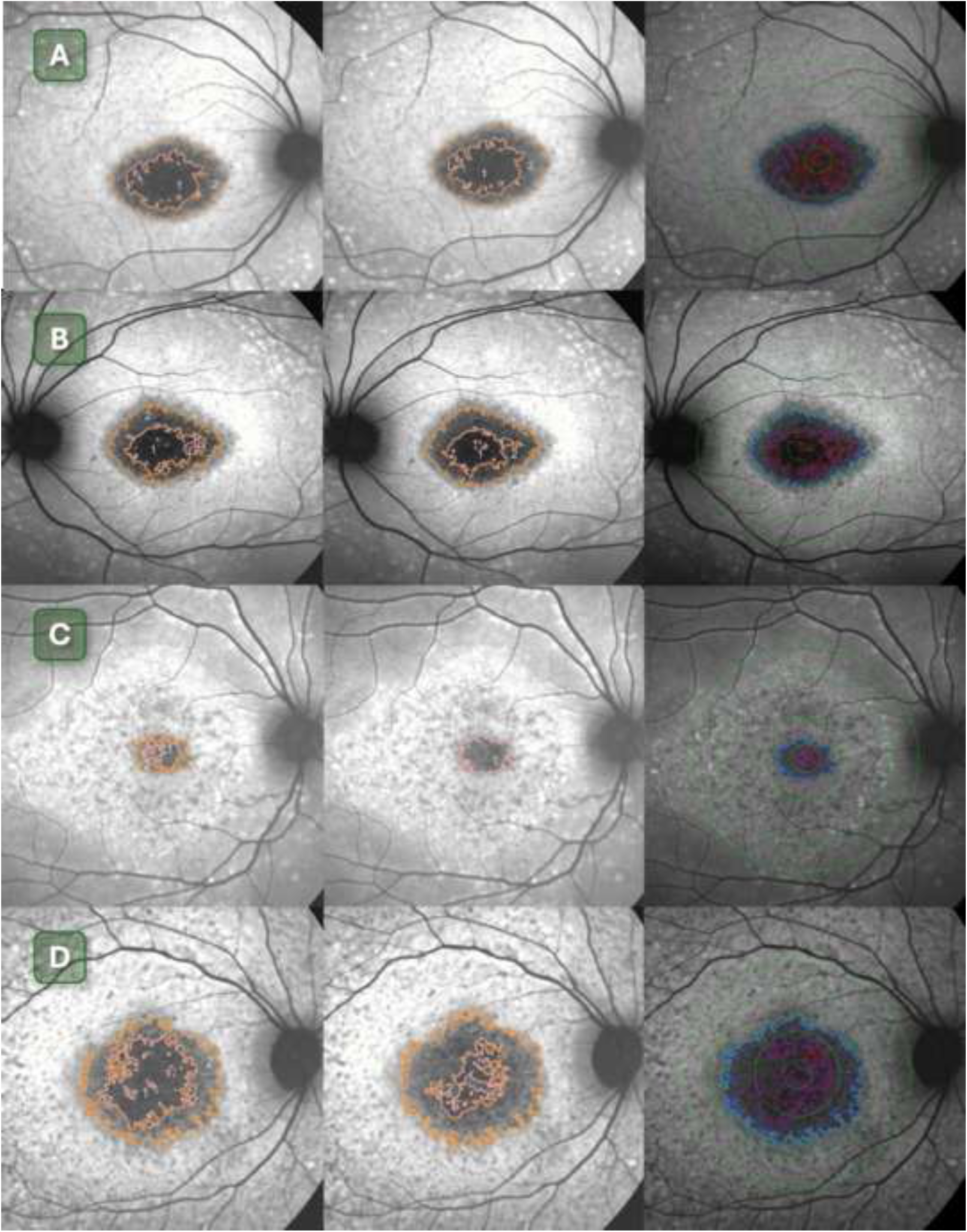
Comparison of manual and automated delineation of decreased autofluorescence (DAF) and definitely decreased autofluorescence (DDAF) lesions on fundus autofluorescence (FAF) imaging in four representative eyes with Stargardt disease. Each row (Panels A–D, top to bottom) represents a different subject. For each, FAF images show lesion segmentation by two representative manual graders (Grader 1, left; Grader 2, center) and one representative automated grader (right). In manual delineations, DDAF regions are outlined in pink and DAF regions are outlined in orange. In automated segmentations, DDAF is defined using a >90% darkness threshold relative to the optic disc (fuchsia), and DAF is defined using a >75% threshold (dark blue). Additional contours reflect alternative thresholds used for visualization only: >70% (light blue) and >95% (red), not included in analysis. Green concentric circles represent ETDRS-like reference zones. **Panel A**: Manual graders show close agreement for both DAF and DDAF, with minor differences in boundary irregularity. The automated delineation mirrors both graders, with slightly larger nasal DDAF extension. **Panel B**: Manual graders agree on overall DAF extent, with Grader 1 identifying a slightly larger DDAF area. The automated segmentation closely matches Grader 1 for both DAF and DDAF. **Panel C**: Both manual graders agree on the presence and extent of DAF, but only Grader 1 identifies a DDAF lesion. The automated segmentation aligns with both graders for DAF and with Grader 1 regarding the presence and boundaries of DDAF. **Panel D**: A large central DAF lesion is consistently delineated by both manual graders, though there is marked disagreement regarding the extent of DDAF. The automated segmentation corresponds well with both graders for DAF and identifies DDAF boundaries that fall between those defined by the manual graders. These examples highlight the variability in manual grading, particularly for DDAF boundaries, and the potential of automated methods to provide reproducible and standardized lesion quantification. Additional threshold contours may offer future utility for stratified staging or progression modeling.

**Table 2.**
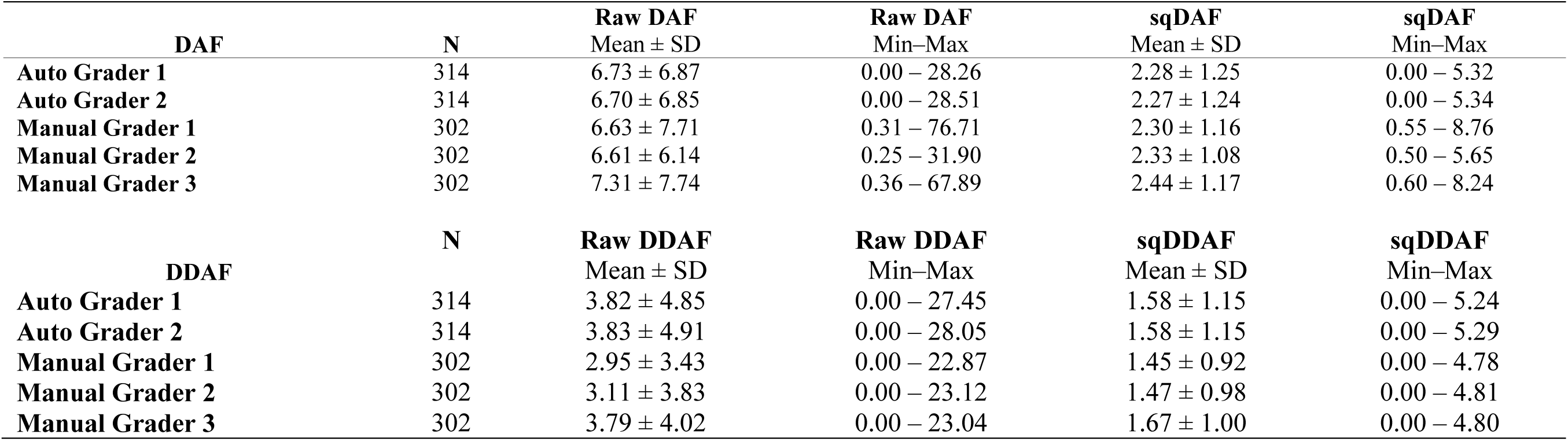
Descriptive statistics for both Decreased Autofluorescence (DAF) and Definitely Decreased Autofluorescence (DDAF) measurements across five graders (two for automated and three for Manual). For each endpoint, both raw lesion areas (in mm²) and their square-root transformed values (sqDAF and sqDDAF) are reported to support statistical analysis and comparability across methods.

### Intra-Grader Repeatability

Automated measurements demonstrated excellent intra-grader repeatability for both DAF and DDAF across graders G1 and G2, with ICCs approaching 0.99 in both raw and transformed data. For example, G1 achieved ICCs of 0.988 (DAF) and 0.989 (DDAF), with corresponding SEM values of 0.69 mm² and 0.44 mm², and MDC/CR values of 1.90 mm² and 1.23 mm², respectively. These metrics remained stable after square-root transformation; the observed reductions in SEM and MDC/CR primarily reflect the compressed measurement scale rather than an intrinsic improvement in repeatability. G2 showed nearly identical results, reinforcing the reproducibility of the automated approach across both lesion types and indicating minimal operator-related variability.

Manual graders also achieved generally high repeatability, particularly for DAF, though variability was greater than in the automated method and differed by lesion type and grader. For DAF, ICCs ranged from 0.909 (G1) to 0.974 (G3) in raw data. While transformation consistently reduced the absolute SEM and MDC/CR values due to scale compression, the degree to which it improved relative measurement precision varied across graders. G1 showed the highest SEM and MDC/CR in raw DAF (2.41 and 6.69 mm²), with notable reduction after transformation (0.28 and 0.78 mm²), though still higher than other graders. G2 and G3 exhibited similar improvements post-transformation, with already stronger baseline performance.

Manual DDAF measurements were less consistent. G1’s ICC improved from 0.837 (raw) to 0.940 (transformed), reflecting heteroscedasticity in lesion size distribution, with corresponding reductions in SEM and MDC/CR. G2 maintained high reliability for DDAF in both scales (ICC 0.947 raw; 0.948 transformed), while G3 showed weaker repeatability, with ICC decreasing from 0.842 to 0.816 after transformation. In G3’s case, the transformation did not confer benefit and may have exposed underlying inconsistencies, as SEM and MDC/CR remained relatively high (1.77 and 4.91 mm² raw; 0.49 and 1.35 mm² transformed).

Overall, intra-grader ICCs ranged from 0.837 to 0.990 in raw data and improved further after transformation (0.816–0.993). The transformation generally reduced measurement error and improved reliability across most graders and endpoints, particularly for manual measurements, although its effect was not uniformly beneficial. Complete metrics are provided in **Table 3**.

**Table 3.**
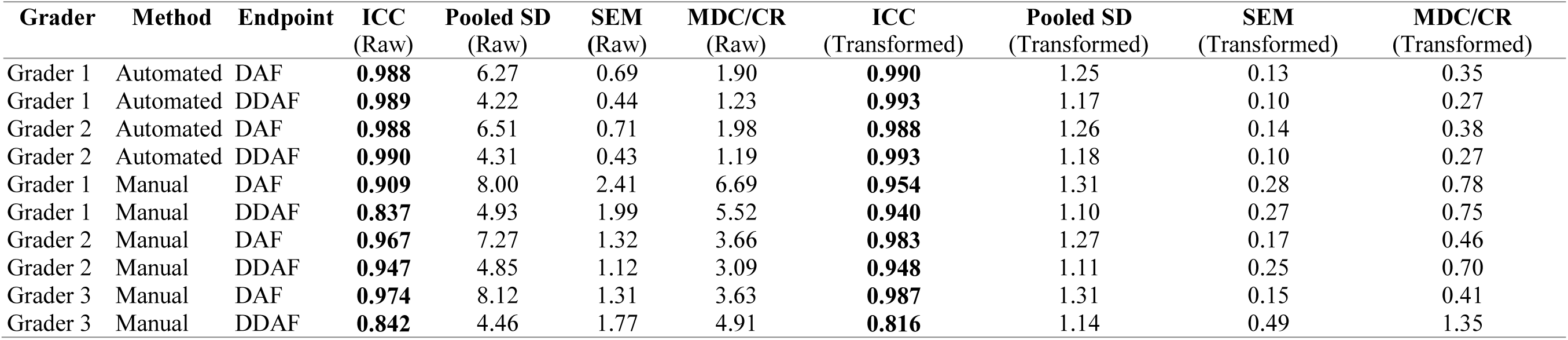
Intra-grader repeatability metrics for Decreased Autofluorescence (DAF) and Definitely Decreased Autofluorescence (DDAF) measurements using raw and square-root-transformed data. The table summarizes intra-grader reliability for three graders (G1–G3) using either manual or automated methods. For each grader–endpoint combination, the table presents the intraclass correlation coefficient (ICC), pooled standard deviation (SD), standard error of measurement (SEM), and minimum detectable change or coefficient of repeatability (MDC/CR), calculated separately for raw and square-root-transformed scales. Higher ICC values and lower SEM/MDC values indicate greater repeatability. Square-root transformation consistently reduced SEM and MDC across both endpoints, particularly for manual measurements of larger lesions, suggesting improved precision and stability of repeated measurements following transformation.

| Grader | Method | Endpoint | ICC<br>(Raw) | Pooled SD<br>(Raw) | SEM<br>(Raw) | MDC/CR<br>(Raw) | ICC<br>(Transformed) | Pooled SD<br>(Transformed) | SEM<br>(Transformed) | MDC/CR<br>(Transformed) |
| --- | --- | --- | --- | --- | --- | --- | --- | --- | --- | --- |
| Grader 1 | Automated | DAF | **0.988** | 6.27 | 0.69 | 1.90 | **0.990** | 1.25 | 0.13 | 0.35 |
| Grader 1 | Automated | DDAF | **0.989** | 4.22 | 0.44 | 1.23 | **0.993** | 1.17 | 0.10 | 0.27 |
| Grader 2 | Automated | DAF | **0.988** | 6.51 | 0.71 | 1.98 | **0.988** | 1.26 | 0.14 | 0.38 |
| Grader 2 | Automated | DDAF | **0.990** | 4.31 | 0.43 | 1.19 | **0.993** | 1.18 | 0.10 | 0.27 |
| Grader 1 | Manual | DAF | **0.909** | 8.00 | 2.41 | 6.69 | **0.954** | 1.31 | 0.28 | 0.78 |
| Grader 1 | Manual | DDAF | **0.837** | 4.93 | 1.99 | 5.52 | **0.940** | 1.10 | 0.27 | 0.75 |
| Grader 2 | Manual | DAF | **0.967** | 7.27 | 1.32 | 3.66 | **0.983** | 1.27 | 0.17 | 0.46 |
| Grader 2 | Manual | DDAF | **0.947** | 4.85 | 1.12 | 3.09 | **0.948** | 1.11 | 0.25 | 0.70 |
| Grader 3 | Manual | DAF | **0.974** | 8.12 | 1.31 | 3.63 | **0.987** | 1.31 | 0.15 | 0.41 |
| Grader 3 | Manual | DDAF | **0.842** | 4.46 | 1.77 | 4.91 | **0.816** | 1.14 | 0.49 | 1.35 |

These findings were further supported by Bland–Altman plots, which visually confirmed the consistency of repeated measurements within each grader (Supplemental **Figure 2** through Supplemental **Figure 5**). For the automated method, both graders demonstrated minimal bias and narrow limits of agreement in raw and square-root-transformed scales for both DAF and DDAF (Supplemental **Figure 2** and Supplemental **Figure 3**). The spread of differences was small and evenly distributed around the mean, with little evidence of heteroscedasticity. In contrast, manual graders exhibited greater variability in repeated measurements (Supplemental **Figure 4** and Supplemental **Figure 5**), especially in raw DAF and DDAF, where wider limits of agreement and skewed distributions were apparent, most notably in Grader 1’s raw DAF and Grader 3’s raw DDAF plots. Square-root transformation improved the agreement patterns for most manual graders by reducing outlier influence and narrowing the limits of agreement, though some grader-specific inconsistencies persisted. Together, these plots reinforce the numerical reliability metrics and highlight the enhanced reproducibility of the automated approach, as well as the benefit, but not guarantee, of variance stabilization via transformation in manual assessments.

**Figure 2.**
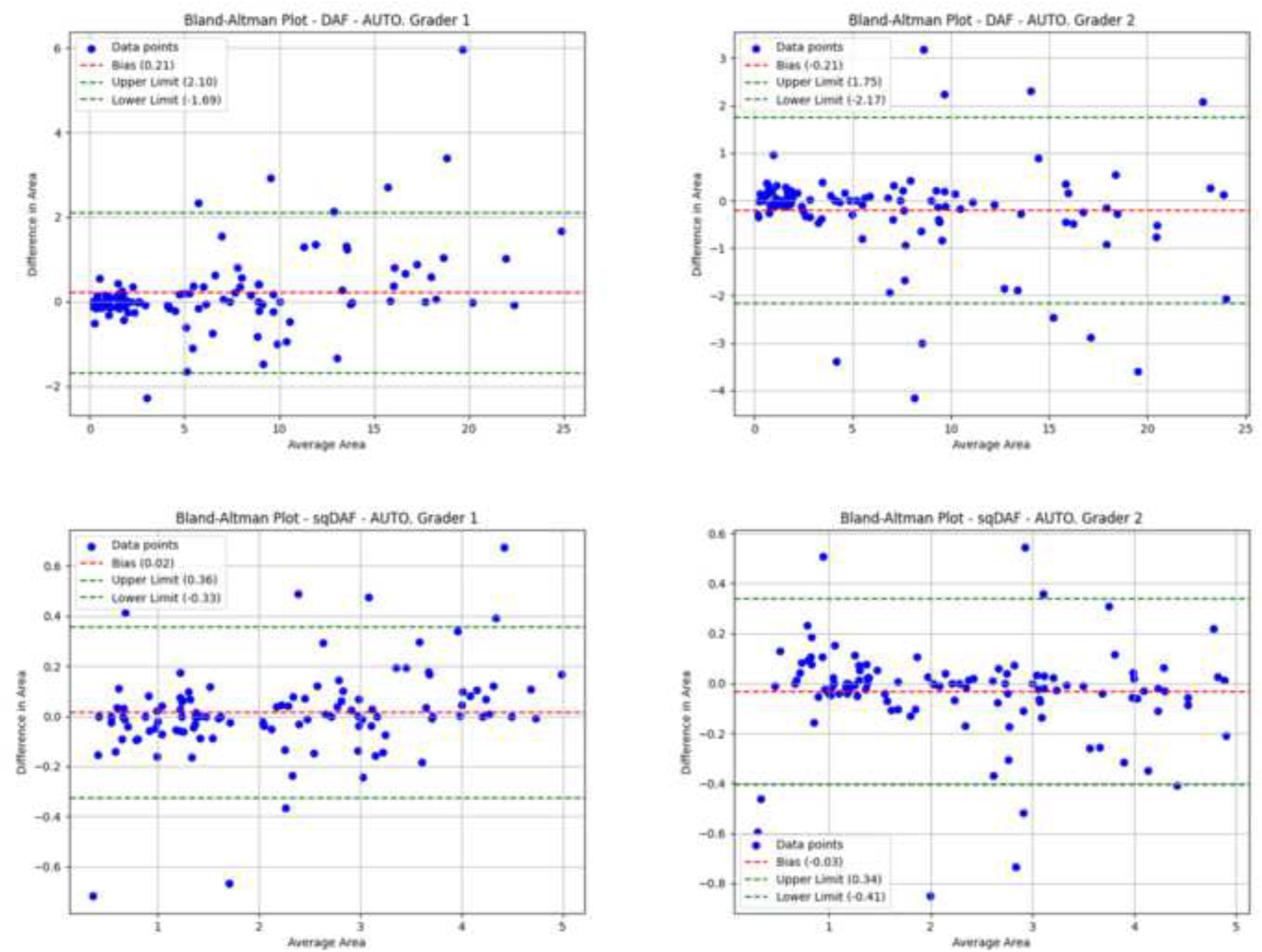
Intra-grader repeatability for Decreased Autofluorescence (DAF) – Automated graders. Bland–Altman plots for raw (top row) and square-root-transformed (bottom row) DAF measurements from two automated graders (left: Grader 1, right: Grader 2). The plots show minimal bias, tight limits of agreement (LoA), and no clear trend of heteroscedasticity, indicating excellent repeatability of the automated method.

**Figure 3.**
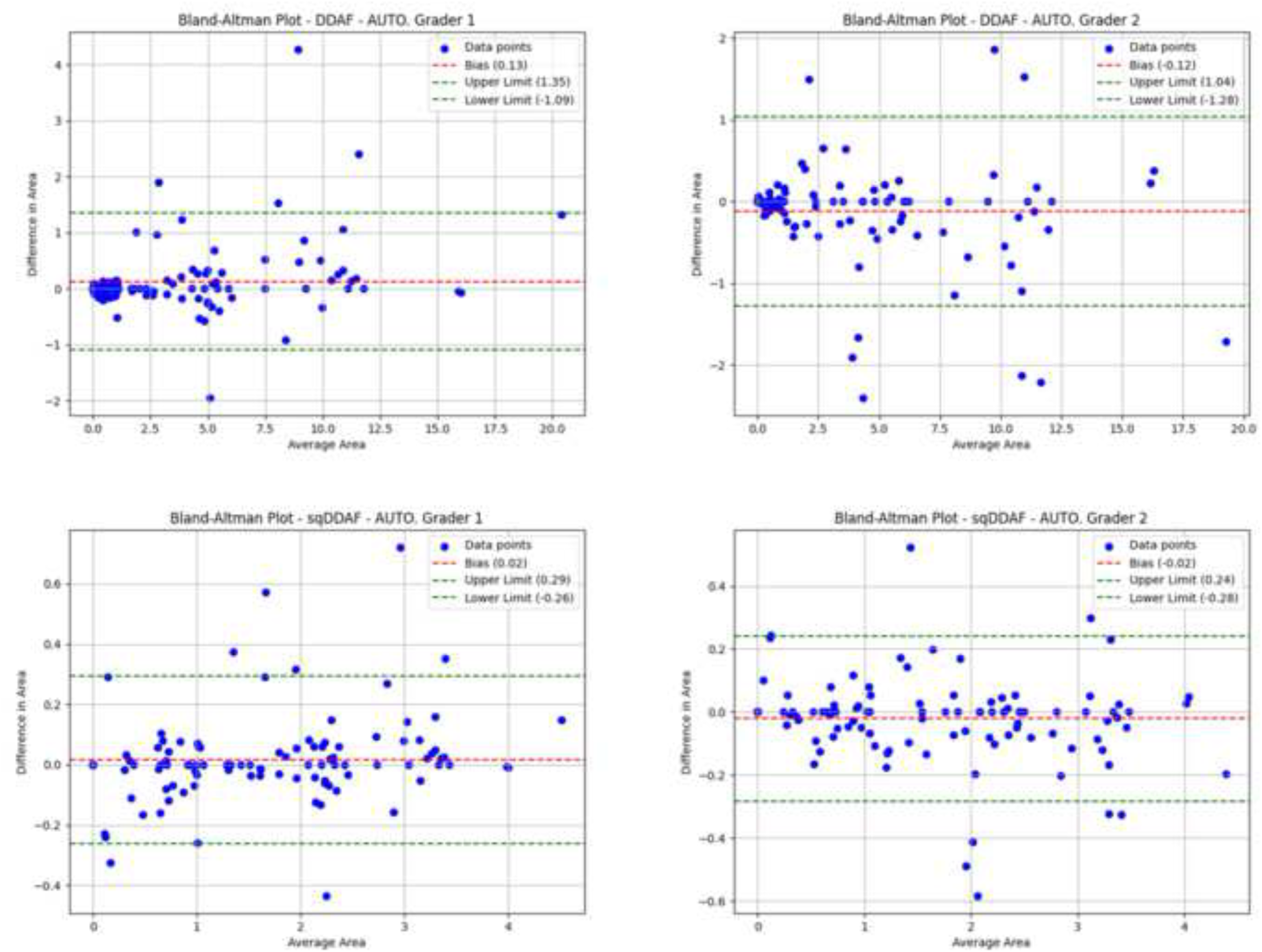
Intra-grader repeatability for Definitely Decreased Autofluorescence (DDAF) – Automated graders. Bland–Altman plots for raw (top) and square-root-transformed (bottom) DDAF measurements for automated Grader 1 (left) and Grader 2 (right). Both sets demonstrate low bias and narrow LoA, with improved uniformity in the transformed plots, reflecting high internal consistency of the automated algorithm.

**Figure 4.**
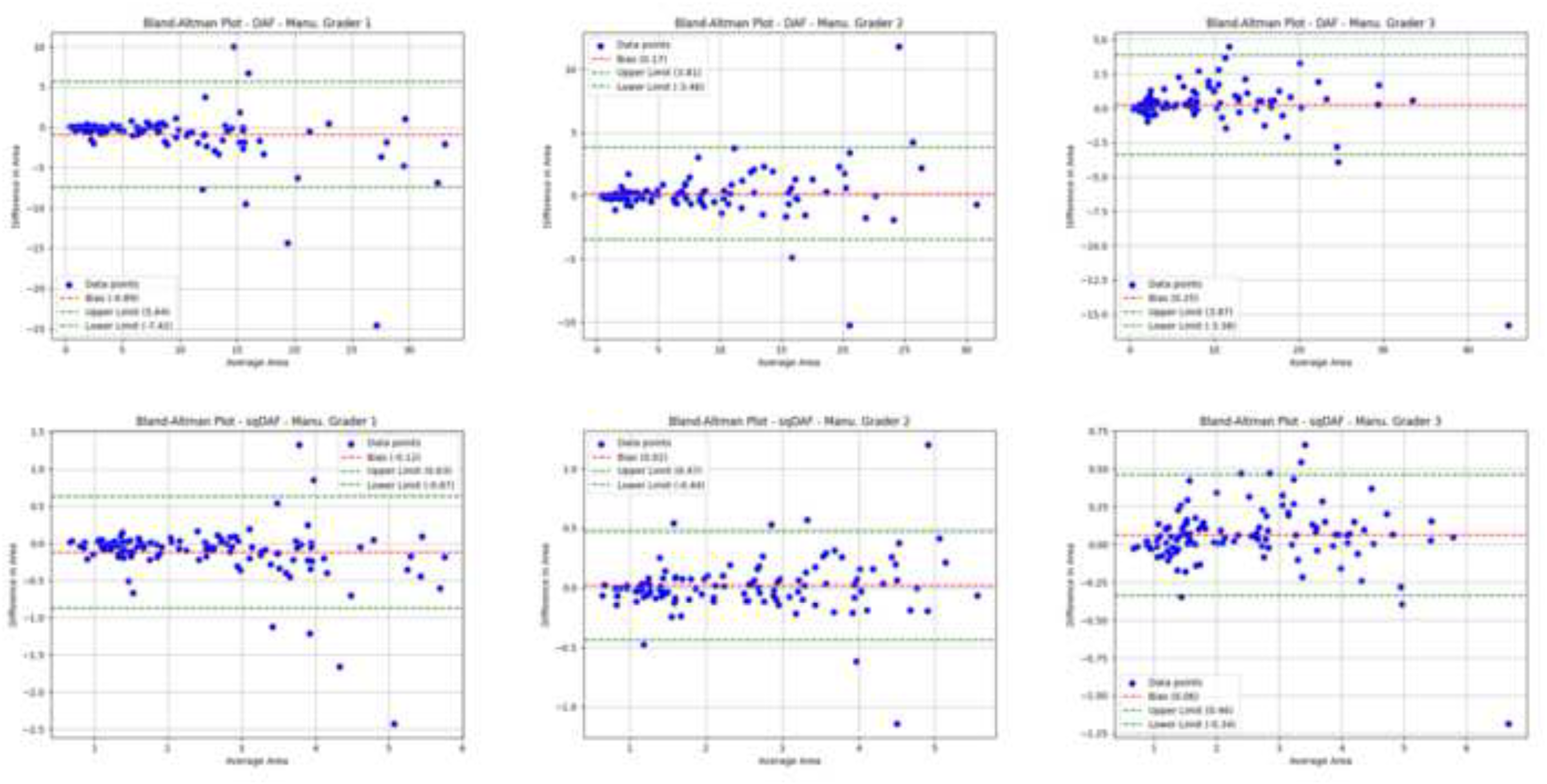
Intra-grader repeatability for Decreased Autofluorescence (DAF) – Manual graders. Bland– Altman plots for raw (top row) and square-root-transformed (bottom row) DAF measurements from three manual graders (left to right: Graders 1–3). While transformation improves uniformity and narrows LoA, raw plots show greater dispersion, especially for larger lesion sizes, indicating variable repeatability across graders.

**Figure 5.**
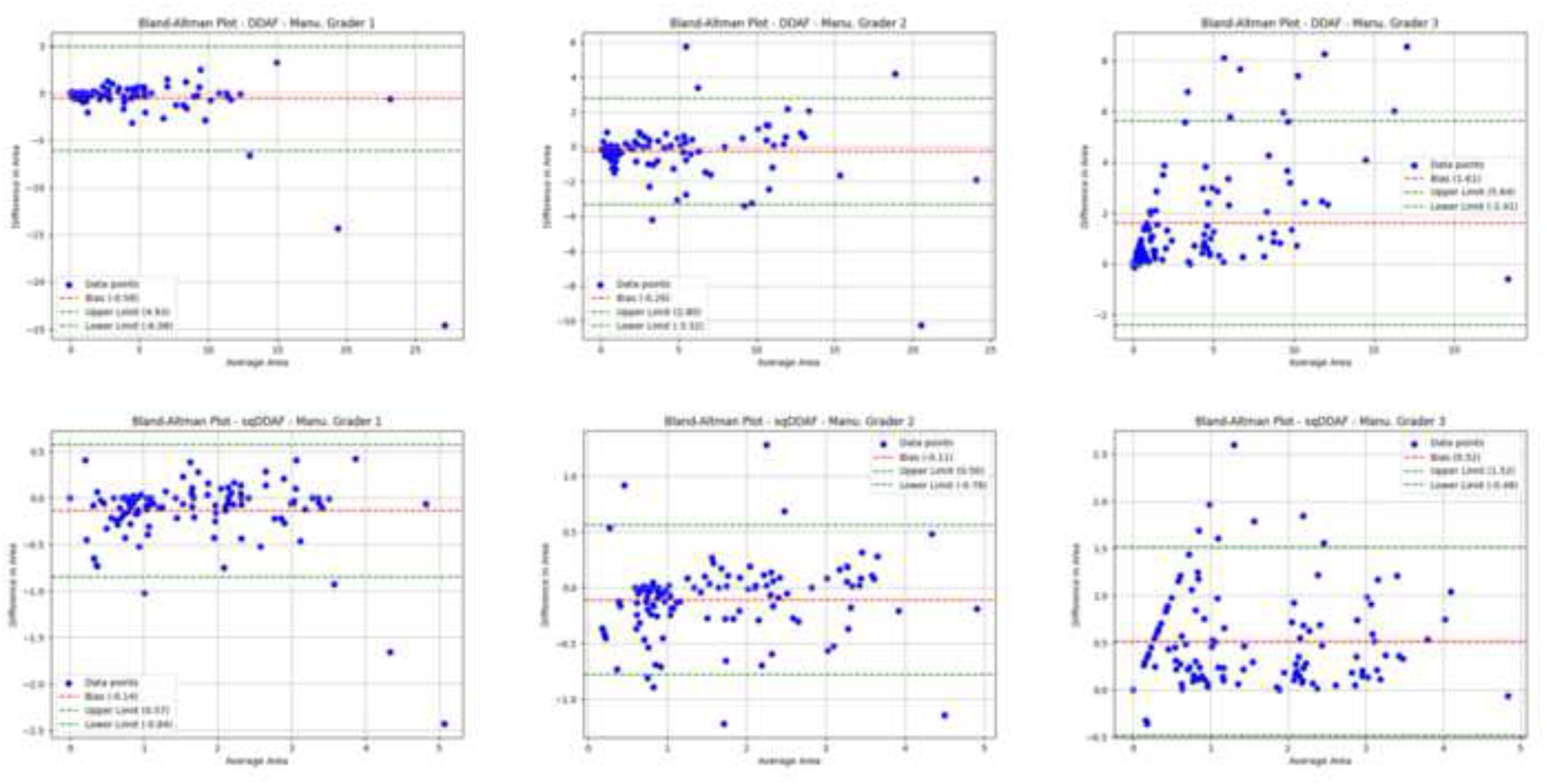
Intra-grader repeatability for Definitely Decreased Autofluorescence (DDAF) – Manual graders. Bland–Altman plots for raw (top) and square-root-transformed (bottom) DDAF from manual Graders 1–3 (left to right). Manual measurements exhibit wider LoA and occasional outliers in raw data; square-root transformation improves agreement for Graders 1 and 2, while variability remains notable for Grader 3.

### Within-Method Reliability (Inter-Grader Repeatability)

Inter-grader repeatability metrics are summarized in **Table 4**, including intraclass correlation coefficients (ICC) for both single- and average-measure reliability (two-way random effects, absolute agreement), along with pooled standard deviation (SD), standard error of measurement (SEM), and minimal detectable change (MDC) at the 95% confidence level.

**Table 4.** Intraclass correlation coefficients (ICCs) for pairwise comparisons of graders on both Decreased Autofluorescence (DAF) and Definitely Decreased Autofluorescence (DDAF), using both raw and square-root transformed measurements. For each comparison, single-measure and average-measure ICC estimates are reported, along with corresponding Standard Error of Measurement (SEM) and Minimal Detectable Change (MDC) at 95% confidence. Values are derived from a two-way random-effects model assuming absolute agreement, reflecting the reliability of individual ratings and the mean of two ratings.

| DAF | Scale | Mean ± SD<br>(Grader 1) | Mean ± SD<br>(Grader 2) | ICC<br>Single (95% CI) | SEM<br>Single | MDC<br>Single | ICC<br>Average (95% CI) | SEM<br>Average | MDC<br>Average | Pooled SD |
| --- | --- | --- | --- | --- | --- | --- | --- | --- | --- | --- |
| Auto. Grader 1 vs. Grader 2 | Raw | 6.73 ± 6.87 | 6.70 ± 6.85 | 0.989 (0.986–0.991) | 0.72 | 1.99 | 0.994 (0.993–0.996) | 0.51 | 1.42 | 6.86 |
| Auto. Grader 1 vs. Grader 2 | sq | 2.28 ± 1.25 | 2.27 ± 1.24 | 0.988 (0.984–0.990) | 0.14 | 0.38 | 0.994 (0.992–0.995) | 0.10 | 0.27 | 1.24 |
| Manual. Grader 1 vs. Grader 2 | Raw | 6.63 ± 7.71 | 6.61 ± 6.14 | 0.764 (0.713–0.807) | 3.38 | 9.38 | 0.866 (0.832–0.893) | 2.55 | 7.06 | 6.97 |
| Manual. Grader 1 vs. Grader 2 | sq | 2.30 ± 1.16 | 2.33 ± 1.08 | 0.906 (0.883–0.924) | 0.34 | 0.95 | 0.951 (0.938–0.961) | 0.25 | 0.69 | 1.12 |
| Manual. Grader 1 vs. Grader 3 | Raw | 6.63 ± 7.71 | 7.31 ± 7.74 | 0.879 (0.849–0.903) | 2.69 | 7.45 | 0.936 (0.918–0.949) | 1.96 | 5.43 | 7.73 |
| Manual. Grader 1 vs. Grader 3 | sq | 2.30 ± 1.16 | 2.44 ± 1.17 | 0.899 (0.868–0.923) | 0.37 | 1.03 | 0.947 (0.929–0.960) | 0.27 | 0.74 | 1.17 |
| Manual. Grader 2 vs. Grader 3 | Raw | 6.61 ± 6.14 | 7.31 ± 7.74 | 0.808 (0.764–0.845) | 3.06 | 8.49 | 0.894 (0.866–0.916) | 2.28 | 6.31 | 6.99 |
| Manual. Grader 2 vs. Grader 3 | sq | 2.33 ± 1.08 | 2.44 ± 1.17 | 0.909 (0.883–0.928) | 0.34 | 0.94 | 0.952 (0.938–0.963) | 0.25 | 0.68 | 1.13 |
| **DDAF** |  |  |  |  |  |  |  |  |  |  |
| Auto. Grader 1 vs. Grader 2 | Raw | 3.82 ± 4.85 | 3.83 ± 4.91 | 0.988 (0.986–0.991) | 0.54 | 1.48 | 0.994 (0.993–0.995) | 0.38 | 1.05 | 4.88 |
| Auto. Grader 1 vs. Grader 2 | sq | 1.58 ± 1.15 | 1.58 ± 1.15 | 0.992 (0.990–0.993) | 0.10 | 0.29 | 0.996 (0.995–0.997) | 0.07 | 0.20 | 1.15 |
| Manual. Grader 1 vs. Grader 2 | Raw | 2.95 ± 3.43 | 3.11 ± 3.83 | 0.939 (0.923–0.951) | 0.90 | 2.50 | 0.968 (0.960–0.975) | 0.65 | 1.80 | 3.64 |
| Manual. Grader 1 vs. Grader 2 | sq | 1.45 ± 0.92 | 1.47 ± 0.98 | 0.922 (0.903–0.937) | 0.27 | 0.74 | 0.959 (0.949–0.967) | 0.19 | 0.53 | 0.95 |
| Manual. Grader 1 vs. Grader 3 | Raw | 2.95 ± 3.43 | 3.79 ± 4.02 | 0.898 (0.793–0.941) | 1.19 | 3.29 | 0.946 (0.885–0.970) | 0.87 | 2.40 | 3.73 |
| Manual. Grader 1 vs. Grader 3 | sq | 1.45 ± 0.92 | 1.67 ± 1.00 | 0.911 (0.789–0.953) | 0.29 | 0.79 | 0.954 (0.882–0.976) | 0.21 | 0.57 | 0.96 |

For DAF, agreement between the two automated graders (Auto G1 vs. G2) was excellent, with ICCs of 0.989 (raw) and 0.988 (square-root), and average-measure ICCs of 0.994 for both scales (**Table 4**). SEM and MDC values were low (≤ 0.72 mm² and ≤ 1.99 mm², respectively), and further decreased following square-root transformation, indicating minimal measurement error.

Manual grader pairs demonstrated moderate-to-high repeatability, with single-measure ICCs ranging from 0.764 to 0.879 in the raw scale and 0.899 to 0.909 in the square-root scale.

Average-measure ICCs improved accordingly (ranging from 0.866 to 0.936 raw and 0.947–0.952 transformed). Square-root transformation reduced the SEM and MDC values across all manual comparisons due in part to the rescaling effect; however, when paired with improvements in ICCs and Bland–Altman agreement, this suggests a potential enhancement in relative measurement consistency.

Bland–Altman plots (Supplemental **Figure 6**) support these findings. For raw DAF, the automated graders showed near-zero mean difference and narrow limits of agreement (LoA), with few outliers or signs of heteroscedasticity. Manual comparisons displayed broader LoA and greater dispersion at higher lesion sizes. In the square-root-transformed plots, differences appeared more uniform across the range of values. The automated pair retained minimal bias and tight LoA, while manual pairs showed reduced spread and fewer extreme outliers, indicating that transformation helped stabilize measurement variance.

**Figure 6.**
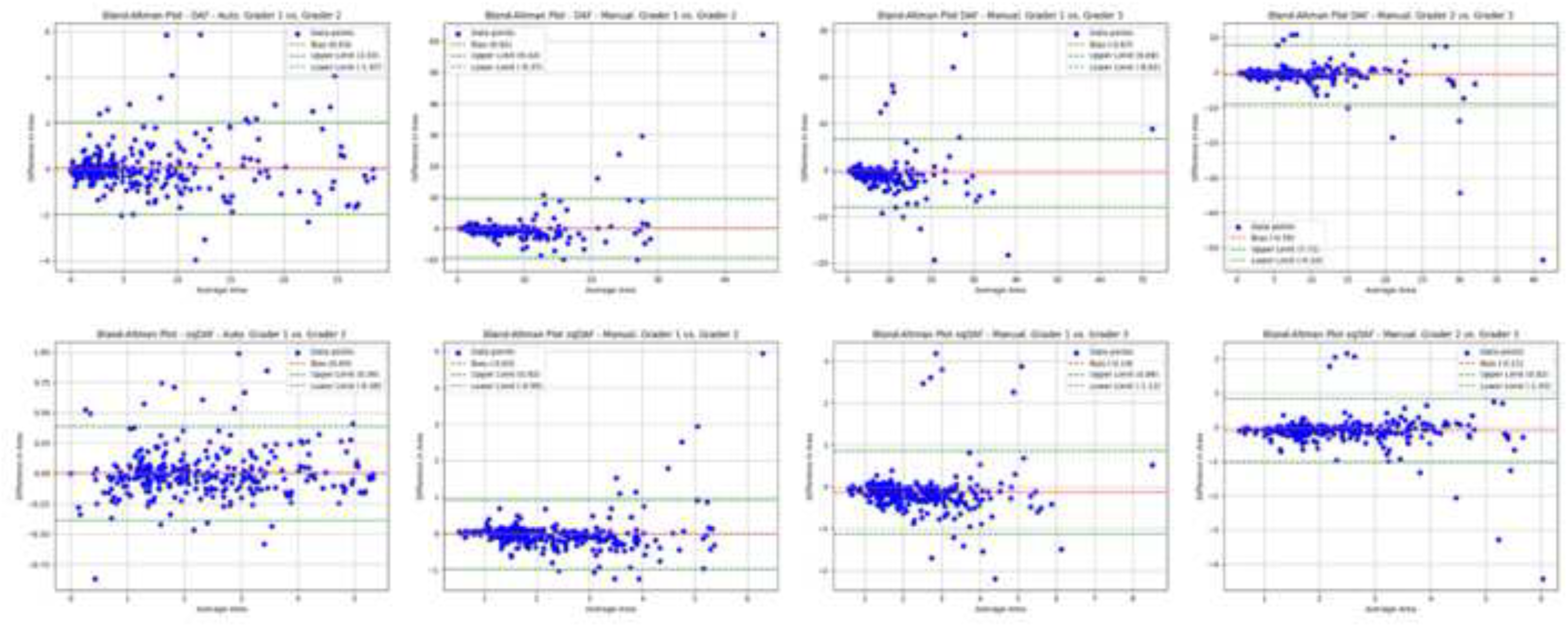
Bland–Altman plots of raw Decreased Autofluorescence (DAF) measurements (top row) and square-root DAF (bottom row), comparing two automated graders (left-most plots) and each pair of manual graders (middle and right plots). The central dashed line indicates the mean difference (bias), while the upper and lower dashed lines represent the 95% limits of agreement (LoA). For the manual comparisons, LoA are visibly wider in the raw scale, particularly at larger average lesion areas, but become narrower following the square-root transformation. The automated graders consistently show minimal bias and narrow LoA in both raw and transformed scales.

Similar trends were observed for DDAF. Automated graders achieved ICCs of 0.988 (raw) and 0.992 (square-root), with average-measure ICCs of 0.994 and 0.996, respectively. SEM and MDC values remained consistently low (e.g., SEM ≤ 0.54 mm²; MDC ≤ 1.48 mm²), reaffirming the method’s precision (**Table 4**). Manual pairs showed high single-measure ICCs (0.892–0.939 raw) and modestly lower values in the transformed scale (0.867–0.922), with average-measure ICCs remaining strong (0.943–0.968 raw; 0.929–0.959 transformed). As with DAF, square-root transformation reduced SEM and MDC values for most manual comparisons due to the scale reduction; in conjunction with improved agreement metrics, this supports its utility for mitigating heteroscedasticity.

Bland–Altman plots for DDAF (Supplemental **Figure 7**) mirrored the DAF pattern. Automated graders exhibited minimal bias and tight LoA in both raw and transformed scales. Manual graders displayed broader LoA in raw data, particularly at larger lesion sizes, but showed visibly reduced dispersion after transformation. These results reinforce that square-root transformation mitigates heteroscedasticity and improves agreement, especially for manual assessments.

**Figure 7.**
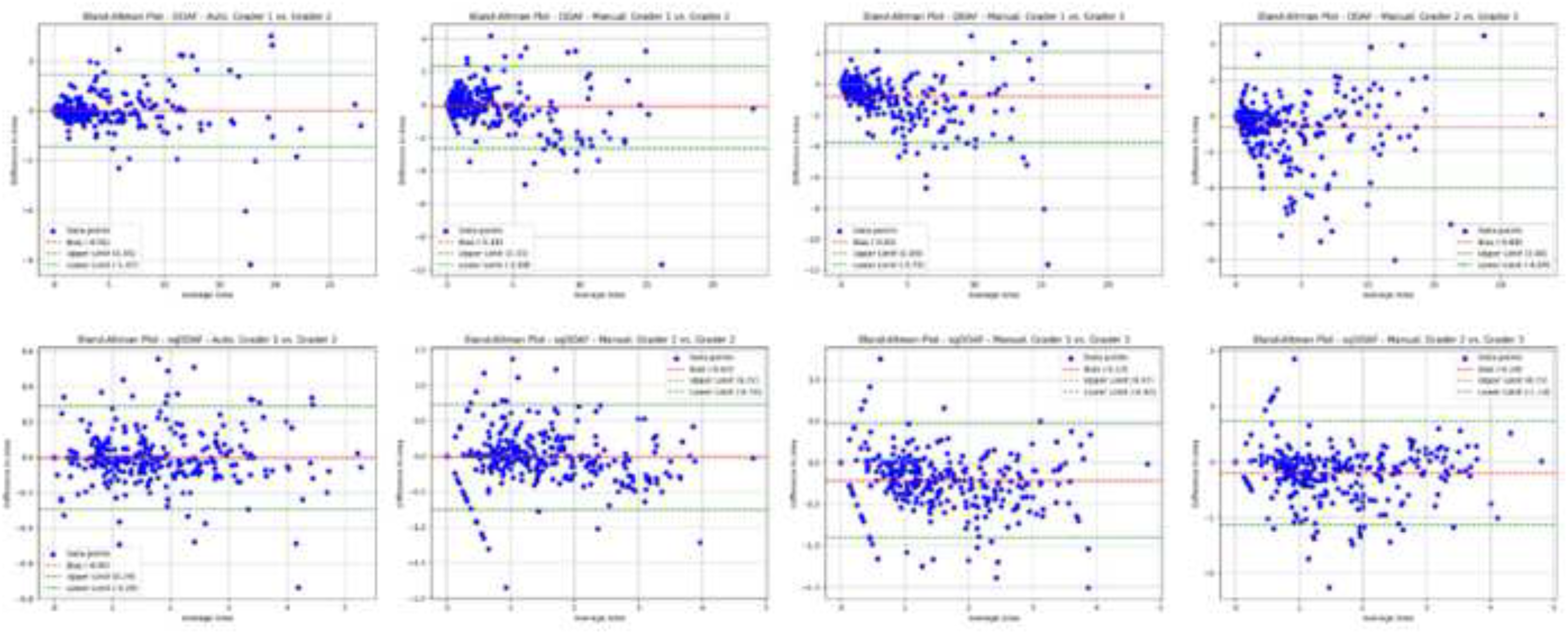
Bland–Altman plots of Definitely Decreased Autofluorescence (DDAF) measurements, showing (top row) raw DDAF for the automated graders (left) and each pair of manual graders (center and right), followed by (bottom row) square-root DDAF. The central dashed lines represent the mean differences (bias), while the upper and lower dashed lines show the 95% limits of agreement (LoA). Automated graders exhibit negligible bias and narrow LoA in both raw and transformed scales, whereas manual graders display greater variability in the raw data, especially at higher lesion sizes, but benefit from noticeably reduced scatter and tighter LoA after square-root transformation.

#### Agreement Analysis (Lin’s CCC and Passing–Bablok)

Agreement between the automated and manual methods was assessed using eyes that were gradable by both approaches and had lesions fully contained within the central 6 mm region of the macula. This constraint is intrinsic to the automated algorithm, which is designed to restrict lesion detection to the central 6 mm area. Inclusion of eyes with lesions extending beyond this region would have introduced a ceiling effect in the automated measurements, potentially distorting the concordance metrics. To ensure valid comparisons, only eyes with lesions measurable by both methods within the same anatomical boundary were included in the analysis. As a result, 14 eyes were excluded, and the final agreement analysis was conducted on data from 288 eyes. Lin’s Concordance Coefficient (CCC) indicated strong agreement for both DAF and DDAF (Supplemental **Figure 8**). In the raw scale, CCC values were approximately 0.91 for DAF and 0.96 for DDAF (**Table 5**), reflecting high concordance between the new automated approach and the manual reference. After square-root transformation, CCC values remained high (0.92 for sqDAF and 0.94 for sqDDAF), confirming that the methods retained strong alignment even when skewness was reduced. Overall, these results reinforce that the automated algorithm’s measurements are largely consistent with manual grading across both lesion endpoints.

**Figure 8.**
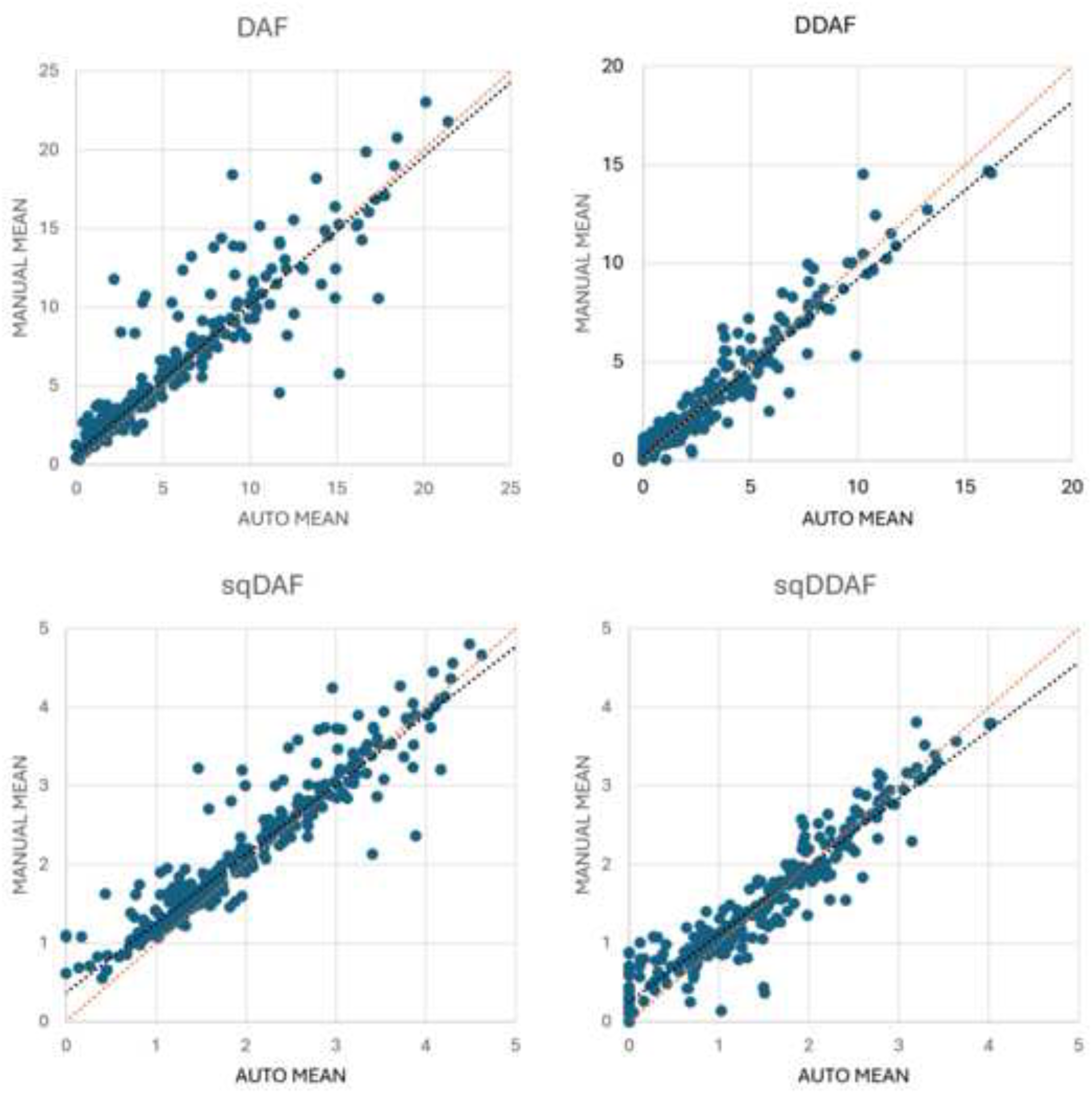
Scatter plots comparing automated and manual mean measurements for Decreased Autofluorescence (DAF) and Definitely Decreased Autofluorescence (DDAF) in both raw and square-root-transformed scales. Each panel displays individual data points (AutoMean vs. ManuMean) for a specific endpoint: raw DAF (top left), raw DDAF (top right), square-root DAF (bottom left), and square-root DDAF (bottom right). The dotted orange line represents the line of identity (Y = X) used in Lin’s Concordance Correlation analysis, while the dotted black line represents the Passing–Bablok regression line fitted to the same data. Visual agreement is strongest where both lines are closely aligned and data points cluster tightly along them. The modest divergence of the Passing–Bablok line from the identity line in some panels reflects slight proportional or constant bias, particularly in raw DAF and sqDAF.

**Table 5.** Summary of agreement analyses between Automated (AutoMean) and Manual (ManuMean) graders for Decreased Autofluorescence (DAF) and Definitely Decreased Autofluorescence (DDAF) (raw and square-root scales). Lin’s CCC quantifies the overall concordance, while Passing–Bablok regression evaluates potential constant or proportional bias between the two methods. 1. Lin’s CCC is computed using the formula ((2ρσ_x σ_y)/(σ_x^2+σ_y^2+(μ_x-μ_y)^2)), where(ρ) is Pearson’s correlation, (σ_x,σ_y) are SDs, and (μ_x,μ_y) are means. 2. Passing–Bablok Slope: Values close to 1 suggest minimal proportional bias; values significantly under or above 1 imply under-/overestimation by one method relative to the other. 3. Passing–Bablok Intercept: Values close to 0 suggest minimal constant bias; positive or negative intercepts point to a fixed offset between the two methods.

| Endpoint | Scale | Lin's CCC | Passing-Bablok | Passing-Bablok |
| --- | --- | --- | --- | --- |
|  |  |  | Slope (median) | Intercept (median) |
| DAF | Raw | 0.91 | 0.945 | 0.665 |
| DAF | sq | 0.92 | 0.880 | 0.369 |
| DDAF | Raw | 0.96 | 0.91 | 0.23 |
| DDAF | sq | 0.94 | 0.864 | 0.247 |

Passing–Bablok regression (Supplemental **Figure 8**) revealed modest intercepts and slopes close to 1 across all comparisons, indicating limited systematic bias. For raw DAF, the slope was 0.945 with an intercept of 0.665, while for sqDAF it was 0.880 with a lower intercept of 0.369 (**Table 5**), indicating minor proportional and constant deviations, respectively. Similar trends were observed for DDAF (Supplemental **Figure 8**): the raw scale showed a slope of 0.910 and an intercept of 0.230, while the square-root scale yielded a slope of 0.864 and an intercept of 0.247 (**Table 5**). These parameters suggest slightly stronger alignment in the raw data but no indication of clinically meaningful deviation in either scale.

## Discussion

This study introduces and validates a novel automated method for quantifying DAF and DDAF in Stargardt disease, demonstrating highly reproducible measurements with minimal grader dependency. ICCs near 0.99 and consistently low SEM and MDC values across graders and lesion types underscore the novel method’s robustness. These results support the use of automated methods in both clinical and research settings where consistency, speed, and scalability are paramount.

In contrast, manual measurements, though generally reliable for DAF, exhibited greater intra- and inter-grader variability, especially for DDAF. This variability likely stems from the inherent difficulty of delineating lesion borders manually, particularly for smaller or irregular DDAF lesions. The variability was most pronounced in Grader 3’s assessments of DDAF, where transformation failed to substantially improve reliability, possibly reflecting systematic grading inconsistencies.

Square-root transformation substantially improved repeatability for most manual measurements by reducing heteroscedasticity. This effect was consistent across graders and endpoints, lowering SEM and MDC and raising ICCs, although not universally. Although square-root transformation reduced SEM and MDC values, these reductions must be interpreted relative to the transformed scale. Improvements in ICCs and agreement plots offer stronger evidence of genuine gains in measurement precision. The transformation had less benefit when systematic bias or grader inconsistency was present, as seen in Grader 3’s manual DDAF data. These findings suggest that transformation is useful for mitigating random variability, but may not fully correct for systematic subjectivity. The difference in reproducibility between DAF and DDAF further highlights the challenges of lesion delineation. DAF measurements were generally more consistent, likely because they are larger, better defined due to better contrast between the lesion boundaries and the normal retinal background, and less susceptible to interpretation bias. In contrast, DDAF lesions in this young cohort were often smaller, patchy, and less well demarcated against a background of QDAF, where the distinction between areas with gray levels >90% and <90% of the optic disc reference is less obvious, contributing to lower agreement. This contrasts with findings from the ProgStar study, which reported higher reproducibility for DDAF, possibly due to inclusion of older patients with more advanced, consolidated lesions. The younger DRAGON cohort, by design, represented earlier disease stages, in which DDAF lesion margins may not yet be clearly defined, increasing manual grading difficulty.

Despite these challenges, the square-root transformation consistently enhanced the reliability of manual assessments, particularly for DDAF. This underscores its value in reducing the impact of outliers and scale-related variance, improving the utility of manual grading in settings where automation is not available.

Agreement between the automated and manual methods was strong across both lesion types, with Lin’s CCC values exceeding 0.90 in all comparisons. Although Passing–Bablok regression revealed mild proportional and constant bias, the deviations were not clinically significant. The automated method’s use of fixed thresholds (e.g., ≥75% of optic disc darkness) may have contributed to slightly elevated lesion size estimates, particularly in DAF. Nonetheless, these small differences are likely correctable through minor calibration if tighter alignment with manual reference standards is desired.

In addition to its measurement precision, the automated method demonstrated greater robustness. Only two images (0.6%) were ungradable by the automated method, compared to 14 (4.4%) by manual grading. The automated approach also yielded fewer outlier exclusions, supporting its capacity to handle image variability and enhance data inclusion in large-scale studies.

Taken together, these findings have important implications. Automated methods offer a consistent and scalable solution for lesion quantification and may significantly reduce intergrader variability in clinical trials. For centers relying on manual grading, applying square-root transformation can improve reliability, particularly for smaller or less defined lesions. However, manual assessment of DDAF remains more variable, underscoring the importance of adopting automated approaches when available.

### Limitations

This study has several limitations. First, its retrospective design inherently restricted the study cohort to the demographic and clinical characteristics defined by the original trial’s inclusion and exclusion criteria, such as age at onset, projected visual acuity, and estimated lesion size. As a result, the findings may not be fully generalizable to the broader Stargardt disease population, particularly those with earlier-stage or more advanced phenotypes. Second, the cross-sectional nature of the study precluded assessment of the longitudinal stability or tracking performance of the automated method compared to the manual approach. Finally, although the automated method demonstrated strong performance, it was evaluated within the context of standardized imaging protocols and a controlled trial environment; its applicability in real-world clinical settings with greater variability in image quality and acquisition remains to be established.

## Data Availability

All data produced in the present study are available upon reasonable request to the authors.

